# Use of Noisy Labels as Weak Learners to Identify Incompletely Ascertainable Outcomes: A Feasibility Study with Opioid-Induced Respiratory Depression

**DOI:** 10.1101/2024.01.29.24301963

**Authors:** Alvin D. Jeffery, Daniel Fabbri, Ruth M. Reeves, Michael E. Matheny

**Affiliations:** School of Nursing, Vanderbilt University, Department of Biomedical Informatics, Vanderbilt University Medical Center Tennessee Valley Healthcare System, U.S. Department of Veterans Affairs Nashville, TN, USA; Department of Biomedical Informatics, Vanderbilt University Medical Center Nashville, TN, USA; Department of Biomedical Informatics, Vanderbilt University Medical Center Tennessee Valley Healthcare System, U.S. Department of Veterans Affairs Nashville, TN, USA

## Abstract

**Objective:** Assigning outcome labels to large observational data sets in a timely and accurate manner, particularly when outcomes are rare or not directly ascertainable, remains a significant challenge within biomedical informatics. We examined whether noisy labels generated from subject matter experts’ heuristics using heterogenous data types within a data programming paradigm could provide outcomes labels to a large, observational data set. We chose the clinical condition of opioid-induced respiratory depression for our use case because it is rare, has no administrative codes to easily identify the condition, and typically requires at least some unstructured text to ascertain its presence.

**Materials and Methods:** Using de-identified electronic health records of 52,861 post-operative encounters, we applied a data programming paradigm (implemented in the Snorkel software) for the development of a machine learning classifier for opioid-induced respiratory depression. Our approach included subject matter experts creating 14 labeling functions that served as noisy labels for developing a probabilistic Generative model. We used probabilistic labels from the Generative model as outcome labels for training a Discriminative model on the source data. We evaluated performance of the Discriminative model with a hold-out test set of 599 independently-reviewed patient records.

**Results:** The final Discriminative classification model achieved an accuracy of 0.977, an F1 score of 0.417, a sensitivity of 1.0, and an AUC of 0.988 in the hold-out test set with a prevalence of 0.83% (5/599).

**Discussion:** All of the confirmed Cases were identified by the classifier. For rare outcomes, this finding is encouraging because it reduces the number of manual reviews needed by excluding visits/patients with low probabilities.

**Conclusion:** Application of a data programming paradigm with expert-informed labeling functions might have utility for phenotyping clinical phenomena that are not easily ascertainable from highly-structured data.

## 1. INTRODUCTION

Although researchers now have access to many large biomedical data sets for extracting meaningful insights, assigning outcome labels in a timely, accurate, and scalable manner remains challenging. Further, many outcomes are not directly ascertainable in a straightforward manner, such as those lacking administrative codes or standardized vocabulary mappings.[1,2] There is growing interest within the informatics community in more nuanced outcomes for clinical decision support, targeted risk model development, and other personalized prediction modalities, all of which require data from multiple sources (e.g., laboratory values, prescriptions) in diverse formats (e.g., structured billing codes, unstructured notes). Leveraging these rich resources for precision health requires a flexible design that permits multiple levels of validation, from spot-checking to validating development iterations to full systematic performance evaluation.

Manual (human) review of records has traditionally been considered the gold-standard of phenotyping. Manual reviews are time-consuming and resource-intensive, particularly within extremely large data sets of patient records.[3,4] To overcome this challenge within electronic health records (EHRs), two broad approaches have been used: (a) condition-specific algorithms that leverage rule-based logic incorporating heterogenous data sources such as diagnostic billing codes, clinical notes, and laboratory values, among others,[5,6] and (b) high-throughput methods that assign thousands of phenotypes to the EHR data, such as PheCodes which are groupings of diagnostic billing codes.[6–9] Condition-specific algorithms are time-consuming to develop/validate, inflexible, and not easily generalizable. Drawbacks of high-throughput methods include: (a) available phenotypes might not include the specific phenotype identified *a priori* by an investigator, and (b) investigators could be interested in less well-defined phenotypes that lack diagnostic billing codes (e.g., adverse events) or where coding practices change for policy reasons (e.g., opioid use disorder-related diagnoses).

To expedite the labeling process, some have advocated for *noisy* labels where investigators train a machine learning model using a large data set with imperfect labels (i.e., some inaccuracies present). One accepts a less stable (i.e., noisy) measure as a tradeoff for the relatively higher expenditure of time and resources needed to procure clean (i.e., high confidence in accuracy) labels from smaller data sets.[10–12] Noisy labels can be derived: (a) from experts creating heuristics that generate an approximation of the ground truth label across all records, or (b) by using a readily-available proxy label that is correlated with the ground truth label. One approach that combines experts’ knowledge with the speed of data-driven approaches is *anchor learning*.[13] In anchor learning, an expert creates rules that serve as an imperfect (or *noisy*) label on which to build supervised models that can generalize beyond the specified anchors and yield the probability of a record having the label of interest. This framework has been applied to healthcare and standardized within the Observational Health Sciences and Informatics (OHDSI) network and as the Automated Phenotype Routine for Observational Definition, Identification, Training, and Evaluation (APHRODITE).[14] A related, newer, and less-evaluated framework is the *data programming* paradigm, introduced by Ratner et al. at Stanford University,[15] that involves specifying multiple imperfect labels developed by experts and has the benefit that phenotypes need not be well-established, universally agreed-upon phenotypes. If we assume each label performs better than chance, one can think of each label as a *weak learner* that has the advantage of being computationally simple while also having the opportunity to be aggregated into an ensemble model that performs better than the sum of its parts.

Respiratory failure is the most common adverse event among perioperative patients (9.13/1000 patients[16]) and costs up to $23.5 billion annually.[17] Surgical patients are particularly susceptible to respiratory depression due to opioid administration for postoperative analgesia. Opioid-induced respiratory depression (OIRD) has an incidence of 0.1-26.9%.[18,19] Operational definitions of OIRD have included naloxone administration, hypoventilation, hypercarbia, and oxygen de-saturation.[18,20] The lack of a standardized definition makes large-scale, observational research challenging due to the difficulties in the assignment of outcome labels. In manual chart reviews, it can be difficult for a reviewer to determine if naloxone administration resulted in the intended benefit. It is not uncommon for naloxone to be administered in the setting of altered mental status to determine whether opioids are responsible. Simply because a patient is receiving opioids, however, does not mean OIRD is the etiology of their altered mental status. To our knowledge, there is no automated approach to identifying OIRD. The most similar criteria would be Patient Safety Indicator (PSI) 11 focused on all-cause post-operative respiratory failure from the Agency for Healthcare Research and Quality (AHRQ).[21,22]

### 1.1. Study Objective

To address these phenotyping challenges, this study examined whether noisy labels generated from subject matter experts’ heuristics using heterogenous data types within a data programming paradigm could be used to provide outcome labels for OIRD within a large, observational dataset.

## 2. METHODS

### 2.1. study design and setting

We conducted a retrospective cohort study using data from the “Synthetic Derivative” database at Vanderbilt University Medical Center, which is a de-identified copy of the main hospital medical records created for research purposes. To perform phenotyping, we used a data programming paradigm (incorporated into the Snorkel software program developed by Ratner et al.[15]) that leverages labeling functions (LFs) as noisy labels to develop a Generative model. The Generative model yields the probability that a record contains the phenotype, which then serves as the outcome in a Discriminative model yielding final labels for the original data set (see Figure 1). We received IRB approval for all activities involving human subjects.

**Figure 1.**
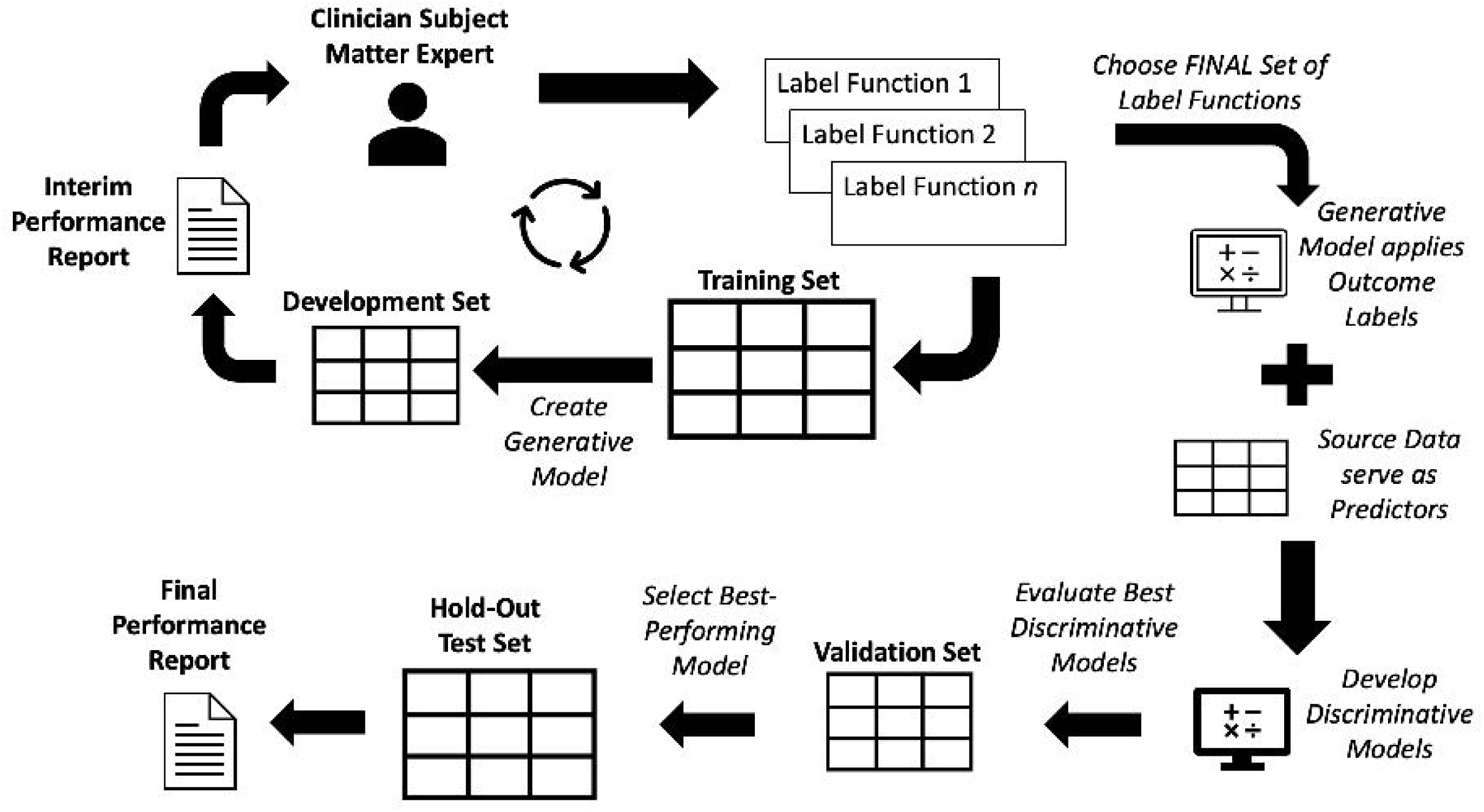
Graphical representation of research methods.

### 2.2. cohort selection

We limited the cohort to surgical procedures included in the AHRQ Patient Safety Indicator-11 (Postoperative Respiratory Failure Rate) based on billing codes.[21,22] Criteria were expected to capture all post-operative OIRD events except for those where prevention is highly unlikely (e.g., those with increased risk for respiratory failure, people with degenerative neurological disorders). As a proxy for elective status (which was not available in our de-identified database), we excluded encounters where the qualifying surgical procedure occurred on the same day as an Emergency Department visit.

Our study cohort comprised 52,861 visits representing 44,999 patients, which we partitioned according to Table 1 (and Figure 2). We first created the Test Set from the visits of patients in the cohort with genetic data (n=2,189 patients), which will be used for a separate study. We included all visits that met AHRQ PSI-11 criteria (n=264, 0.50% of cohort) and randomly sampled 500 visits (0.95% of cohort) that did not meet criteria. Of the remaining 52,097 visits, we excluded 285 visits because they were associated with patients who had visits already included in the Test Set. Then, we randomly selected 50 visits for the Validation Set and Development Set using 2:1 over-sampling with 2 AHRQ-defined cases per 1 AHRQ-defined control. We enriched the Validation and Development Sets throughout the study, as described in the next section.

**Figure 2.**
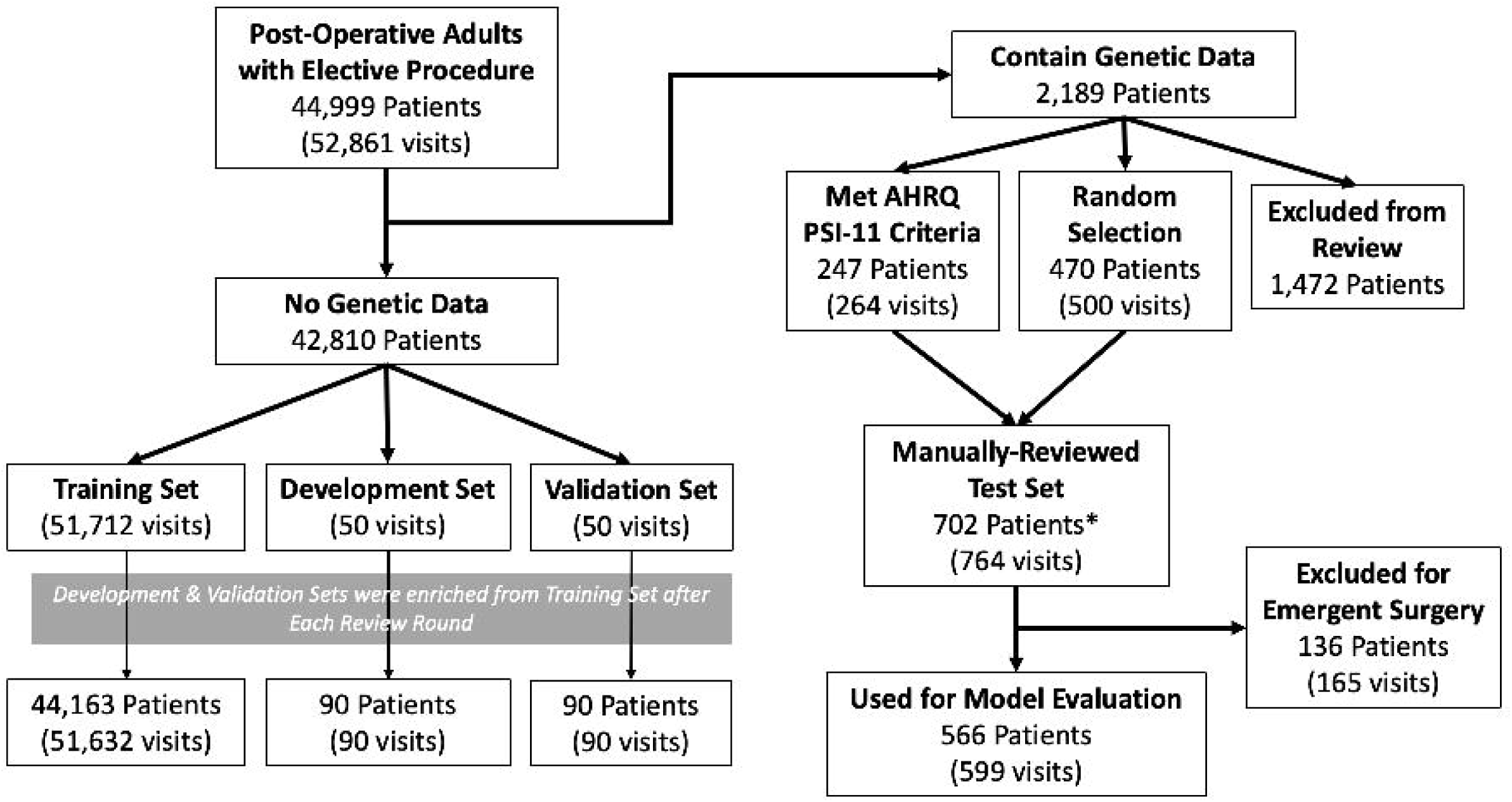
Flow diagram illustrating the number of patients and visits present at each phase of cohort processing. \**Note:* The number of unique patients in the Manually-Reviewed Test Set (702) is smaller than the sum of the 2 preceding boxes (717) because those boxes were sampled at the visit-level instead of patient-level.

**Table 1.**
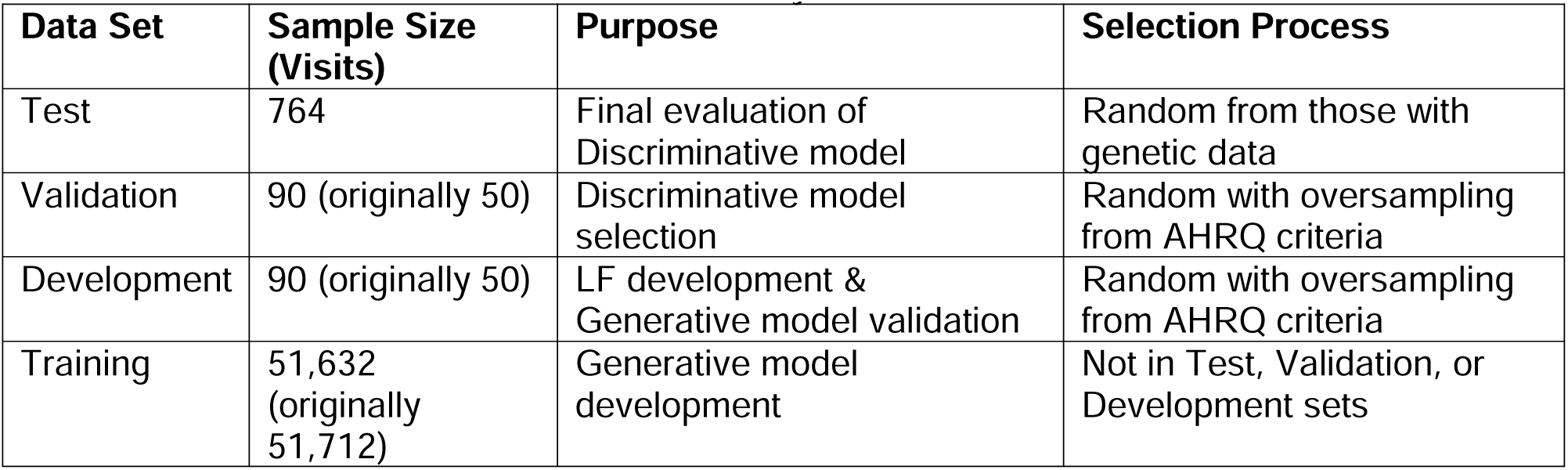
Characteristics of data sub-sets for study.

### 2.3. generative model development and evaluation

Developing the Generative model involved an iterative process of: (a) developing candidate LFs, (b) examining candidate LF performance in the Development Set, (c) using the Python-based Snorkel software to develop a candidate Generative model in the Training Set, and (d) evaluating performance of the candidate Generative model in the Development Set.

#### 2.3.1. Labeling Function Creation

In the data programming paradigm, a developer writes *labeling functions* (LF) that serve as noisy labels based on heuristics, patterns, or external information. Each LF processes input data and returns a vote of a Yes (1), No (0), and/or Abstain (−1). LFs can be overlapping such that multiple LFs use the same input data. LFs can be conflicting such that the same record yields different votes (e.g., one LF yields a Yes vote while another LF yields No vote on the same record). While LFs could potentially return any of the three vote options, each LF only needs to return 2 of the 3 vote options (e.g., Yes versus Abstain, Yes versus No). LFs produce an *m x n* label matrix with *m* examples and *n* LFs. Without any ground-truth data, we can use the label matrix to model accuracies and correlations between LFs to optimize a Generative model that yields probabilistic labels. We abandoned the suggested context hierarchy[15] in favor of treating an entire visit as a single record/exemplar, which resulted in individual LF performance improvement.

The lead subject matter expert, a dually trained biomedical informaticist and critical care nurse (ADJ), conducted chart reviews of Development Set visits to create candidate LFs and determine whether each visit had evidence of OIRD. LFs comprised data from medication information, clinical note text (using regular expressions for words and phrases), and administrative diagnostic and procedure codes.

Elements guiding LF creation and modification included: (a) coverage – the proportion of visits in which the LF could yield a vote, (b) conflicts – whether another LF yielded a different vote, and (c) empirical accuracy – the proportion of visits correctly labeled, excluding Abstain votes, based on the single reviewer’s determination.

#### 2.3.2. Iterative Model Development

We used Snorkel to generate a probability of whether a visit included an OIRD event. Due to the low number of positive Cases in the initial Development Set (2/50, 4%), we applied this iterative process to enrich the Development Set and Validation Set by extracting visits with the top 20 probability values from the Training Set and dividing those equally among the Development Set and Validation Set. After Round 4, the primary LF developer facilitated a focus group with clinicians and biomedical informaticists to discuss face validity of the current LFs and solicit additional heuristics for additional LFs (Table 2).

**Table 2.**
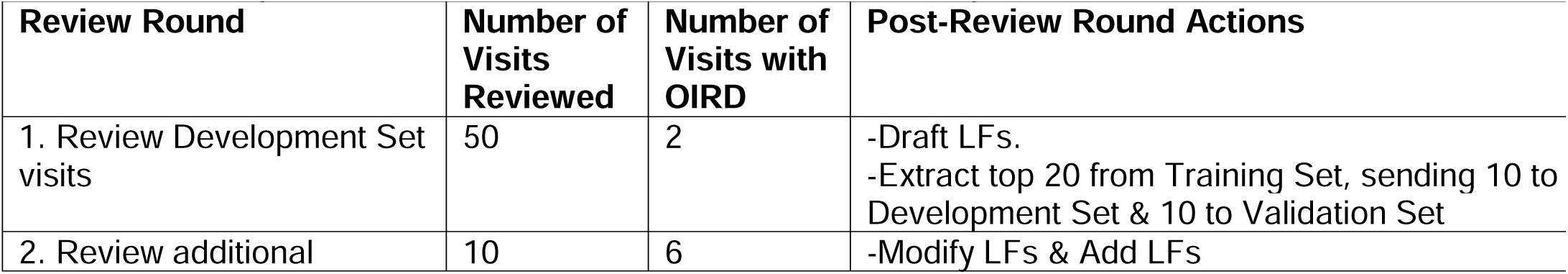
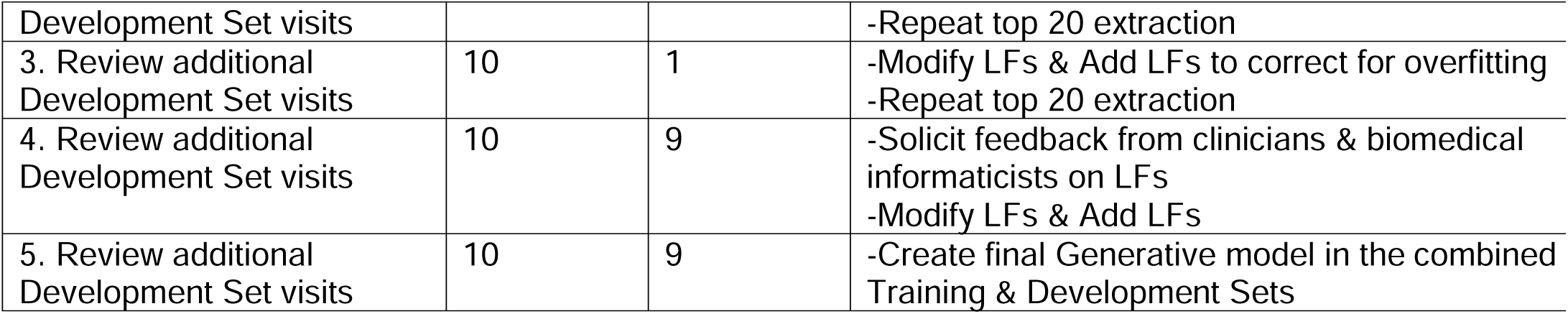
Labeling function (LF) development process with Training and Development Sets.

We conducted hyper-parameter tuning of the Generative Model’s neural networks using learned LF weights in the Training Set and empirical accuracy in the Development Set (except in the final round where we combined the Training Set and Development Set and used Validation Set to assess empirical accuracy). We proposed a new method for Generative model hyper-parameter tuning by emphasizing the learned weights of the LFs rather than focusing on empirical accuracy, a modification which makes theoretical sense but should be examined more robustly in future studies. We selected hyper-parameters that yielded higher LF weights for clinically important rules, which were determined by two clinicians on the research team (a nurse practitioner and a physician). For example, an LF that used information about naloxone administration (i.e., a specific treatment for OIRD reversal) should be more important than an LF that assessed for altered mental status, which is less specific to OIRD.

### 2.4. discriminative model development and evaluation

We used the final Generative model’s probabilistic labels as outcome labels for a Discriminative model. Unlike the Generative model that makes predictions using the output from LFs, the Discriminative model uses features directly from the source data. This step confers the added benefit of increased generalizability. A Discriminative model can be used by external stakeholders or with future unlabeled data without needing the Snorkel software, LFs, or access to the same input data used for the Generative model. Additional benefits of a Discriminative model are: (a) the ability to include the labels from manually-reviewed records to improve the model’s performance and (b) the option to use a noise-aware model to account for uncertainty within the probabilistic Generative labels.

#### 2.4.1. Candidate Variables

We selected age, gender, binary indicators related to administrative codes and naloxone administration, and frequency of keywords/phrases in clinical notes to serve as predictors. Administrative codes included diagnostic and procedure codes related to respiratory failure/disease, prolonged mechanical ventilation, sepsis, cardiovascular disease, and cerebrovascular accidents. Based on input from clinical subject matter experts on the research team as well as focus group members (see 2.3.2.), we developed keywords and phrases related to naloxone administration and its effectiveness, narcotic overdose, absence of pain medications, decreasing or holding opioids, presence of acute events, altered mental status, pinpoint pupils, and hypoxia. Other predictors included number of notes from respiratory therapists and rapid response team mentions.

#### 2.4.2. Model Development

We began Discriminative model development with off-the-shelf[23] machine learning algorithms from Python’s scikit-learn to identify the most promising algorithms for hyper-parameter tuning. Classification algorithms comprised logistic regression, linear discriminant analysis, k-nearest neighbors, decision trees, random forest, Naïve Bayes, and a multilayer perceptron (i.e., neural network).[23] Regression algorithms comprised linear regression, random forest, and a multilayer perceptron.[23] Based on F1-scores, AUC, and mean squared error,[24,25] we chose the random forest and multilayer perceptron algorithms for hyper-parameter tuning[23] in both the classification and regression tasks.

#### 2.4.3. Internal Model Validation

To estimate the model’s future performance in an unbiased manner, we performed nested cross-validation with a manual grid search on the combined Training/Development Set using 3 inner folds and 10 outer folds. The nested cross-validation suggested F1 scores will range 0.6-0.8 for classifiers and 0.4-0.7 for regressors, AUCs will range 0.75-0.9 for classifiers and 0.6-0.8 for regressors, and mean squared errors will range 0.005-0.008 for classifiers and 0.005-0.01 for regressors. The classification algorithms outperformed the regression algorithms, and the random forest classifiers (weighted and unweighted) outperformed the multilayer perceptron classifier. Given some overlapping performance (dependent on hyper-parameter choices), we trained each of the 5 models using the hyper-parameters that most frequently had the highest performance on the outer folds to serve as our best candidate models and evaluated their performance in the Validation Set. The weighted random forest classifier performed best and was designated the final Discriminative model.

#### 2.4.4. Reference Standard Validation

We compared our final model with the hold-out Test Set that was manually adjudicated the Vanderbilt University Medical Center’s Crowdsourcing Core services. The Crowdsourcing Core assists investigators in describing desired outcomes for clinical chart reviews, recruiting and compensating qualified reviewers (known as “workers”), displaying complex clinical data for review, and ensuring sufficient numbers of reviews to make a determination.[26] Workers completed the review in a two tasks, which were completely independent of the investigative team’s activities. In the first task, workers evaluated whether the visit included an elective surgery. Visits without an elective surgery were excluded from further review. In the second task, workers evaluated whether respiratory depression occurred and whether it was likely due to opioid administration.

#### 2.4.5. Sensitivity Analyses

After final assessment of model performance with all *a priori* decisions, we conducted a *post-hoc* sensitivity analysis to measure the influence of some choices made during the final Discriminative model development. We specified the outcome label from the Generative model’s predicted probability for the 51,712 records in the Training Set; however, for the additional 90 records in the Development Set, we specified the outcome label based on the manually adjudicated determination. Additionally, we weighted samples during model fitting based on the absolute value of the Generative model probability’s distance from 0.5. Values closer to 0 represented low certainty (i.e., a random guess) while values closer to 0.5 represented greater certainty. These weights were used to determine the penalty of misclassifications (i.e., mis-classified predictions where the probabilistic outcome label was closer to 0.5 were penalized less than those closer to 0 or 1).

## 3. RESULTS

### 3.1. labeling functions in training and development sets

We finalized our Generative model with 14 LFs (Table 3).

**Table 3.**
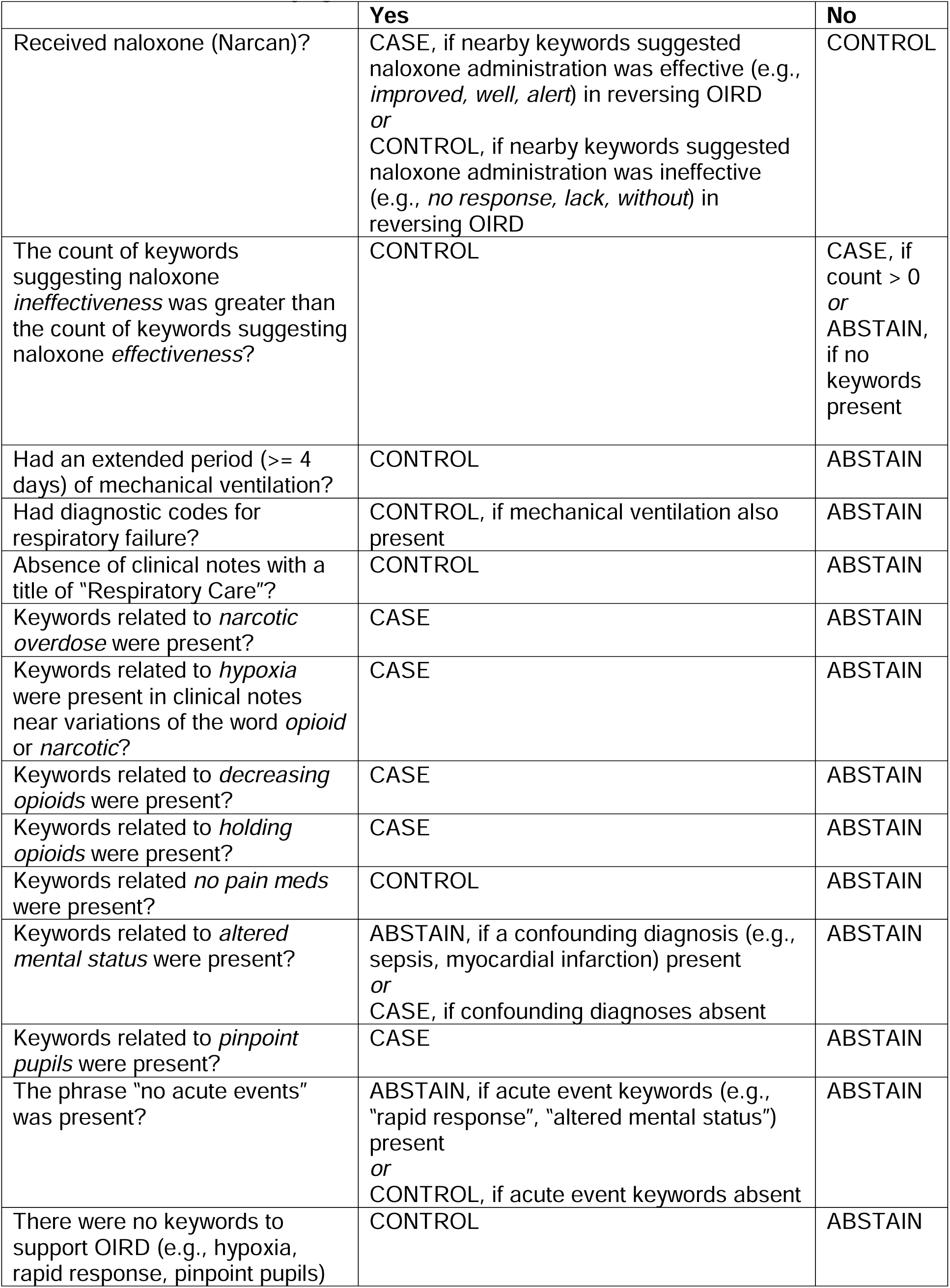
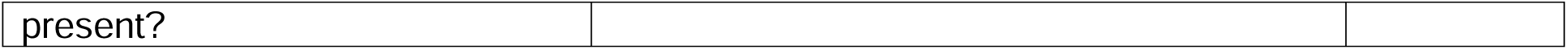
Final LFs for identifying OIRD in the Generative model.

### 3.2. validation set performance

In the Validation Set, the empirical accuracy of individual LFs ranged 0.47-1.00, the final Generative model achieved an accuracy of 0.83, an F1 score of 0.73, and an AUC of 0.96 (Figure 3), and the final Discriminative model achieved an accuracy of 0.88, an F1 score of 0.80, and an AUC of 0.92 (Figure 4). Performance of the final Discriminative model in the Validation Set was consistent with expected performance during internal validation.

**Figure 3.**
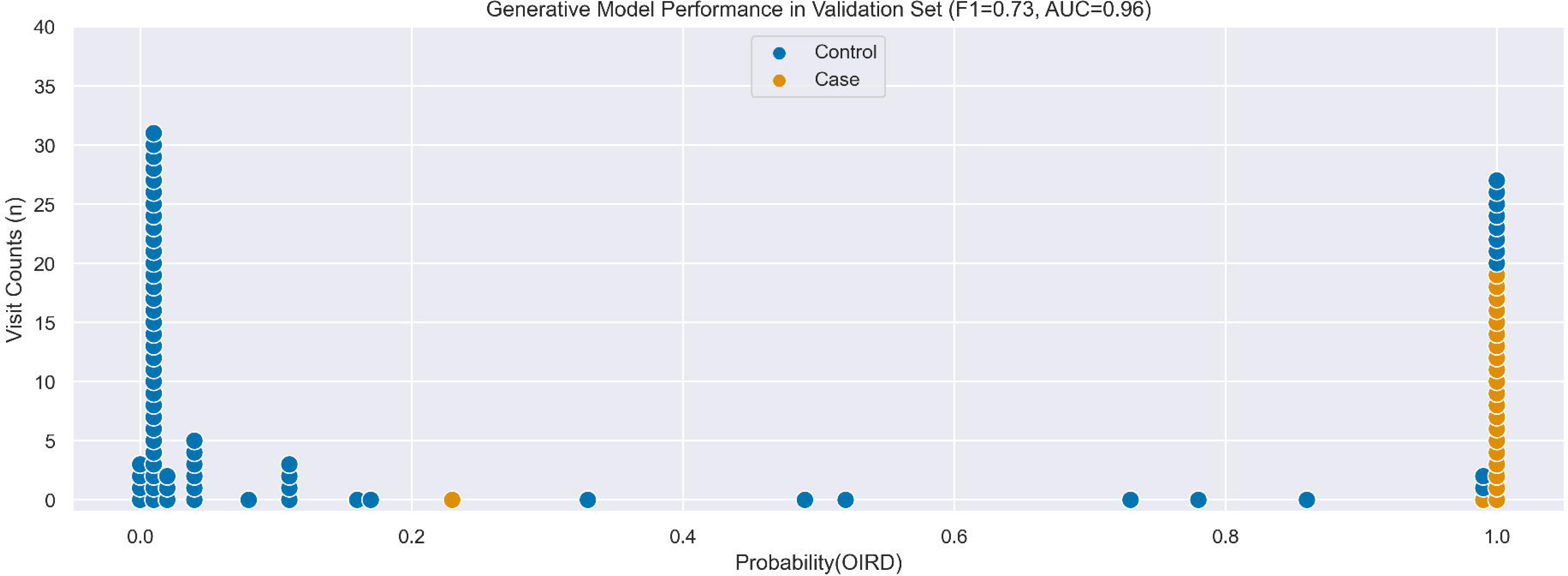
Comparison of OIRD predicted probabilities from the Generative model with manually-adjudicated labels in Validation Set.

**Figure 4.**
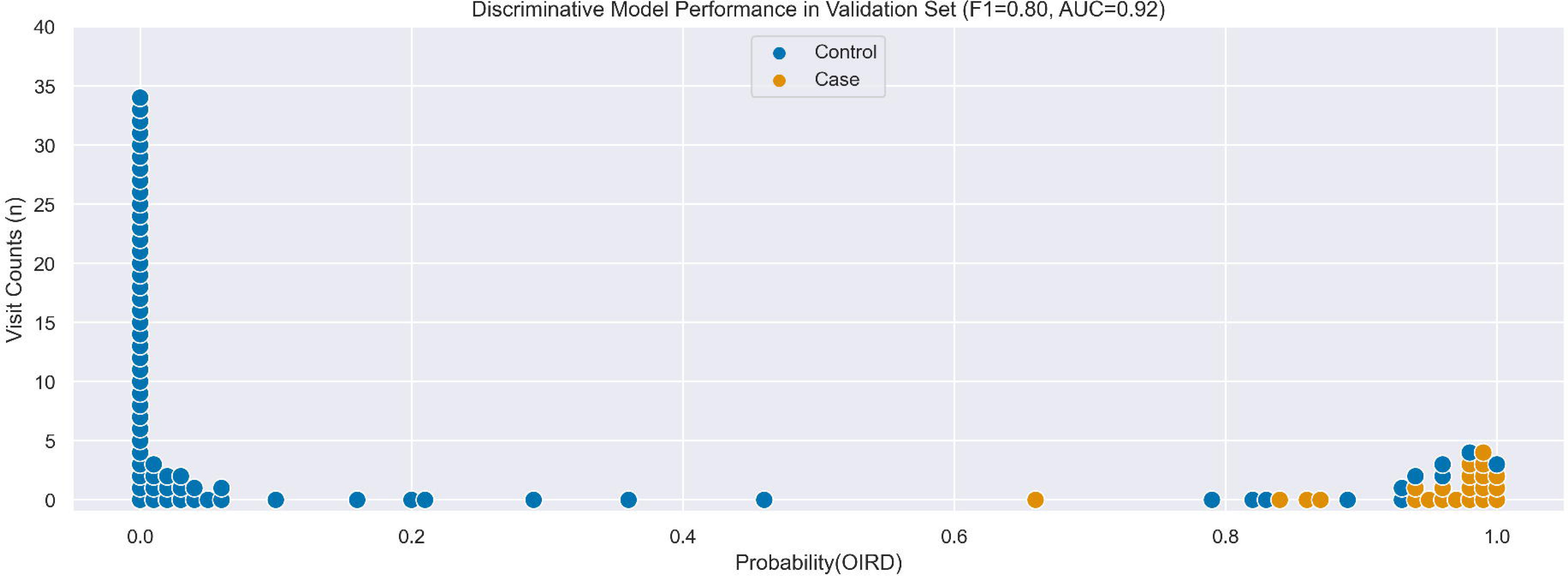
Comparison of OIRD predicted probabilities from the Discriminative model with manually-adjudicated labels in Validation Set.

In the post-hoc sensitivity analysis, the Discriminative model trained with the removal of manually adjudicated outcome labels from the Development Set (i.e., all outcome labels were produced by the Generative model’s probabilistic labels) yielded the same accuracy, F1 score, and AUC values in the Validation Set. Conversely, the Discriminative model trained without sample weighting during the model fit yielded decreased accuracy (0.87), F1 score (0.79), and AUC (0.91) values in the Validation Set. During a review of record-level performance in the Validation Set, records with large a discrepancy between the predicted probabilities of Generative and Discriminative models primarily occurred when the Generative model indicated a probability close to 1 yet the manually adjudicated label was “control.” Therefore, sample weighting during model fit improved overall model performance while the presence of manually adjudicated labels corrected some records mis-classified as being a “case” in the Validation Set data.

### 3.3. test set performance

In the first task, workers excluded 165 visits (21.6%) where the surgery was emergent. In the remaining 599 visits for the second task, workers determined OIRD was present in 5 (0.83%) visits. In the manually adjudicated Test Set, the final Generative *and* Discriminative models achieved an accuracy of 0.977, an F1 score of 0.417, and an AUC of 0.988. A simple majority vote from the 14 LFs resulted in lower accuracy (0.967), F1 score (0.333), and AUC (0.983) values.

The Discriminative models used in the *post-hoc* sensitivity analysis for the Validation Set were associated with improved positive predictive values and F1 scores in the Test Set (Table 4). The original AHRQ criteria performance, which served as a baseline comparison in the Test Set, was lower than the Generative and Discriminative models with an accuracy of 0.677 (vs. 0.977), F1 score of 0.040 (vs. 0.417), and an AUC of 0.738 (vs. 0.988) (Table 4). Using the baseline AHRQ performance, 4 of the original 196 “cases” were determined to be a true case, and 1 of the original 402 “controls” was determined to be a true case.

**Table 4.** Performance of all phenotyping approaches in Test Set.

|  | **Sensitivity** | **Specificity** | **Positive Predictive Value** | **Accuracy** | **AUC** | **F1 Score** |
| --- | --- | --- | --- | --- | --- | --- |
| Majority Vote from Labeling Functions | 1.0 | 0.97 | 0.2 | 0.967 | 0.983 | 0.333 |
| Generative Model | 1.0 | 0.98 | 0.263 | 0.977 | 0.988 | 0.417 |
| Final Discriminative Model (with weighting and some manual labels) | 1.0 | 0.98 | 0.263 | 0.977 | 0.988 | 0.417 |
| Discriminative Model without weighting | 1.0 | 0.98 | 0.278 | 0.978 | 0.989 | 0.435 |
| Discriminative Model without any manual labels | 1.0 | 0.98 | 0.278 | 0.978 | 0.989 | 0.435 |
| Discriminative Model without weighting or any manual labels | 1.0 | 0.98 | 0.278 | 0.978 | 0.989 | 0.435 |
| AHRQ PSI-11 Criteria | 0.8 | 0.68 | 0.020 | 0.677 | 0.738 | 0.040 |

When examining the final Test Set status in the context of both the Generative and Discriminative models, all of those identified as a Case have a Generative model probability > 0.8 and a Discriminative model probability > 0.7 (Figures 5 and 6). If one used these higher, joint thresholds, a revised scoring system would have an F1 score of 0.625.

**Figure 5.**
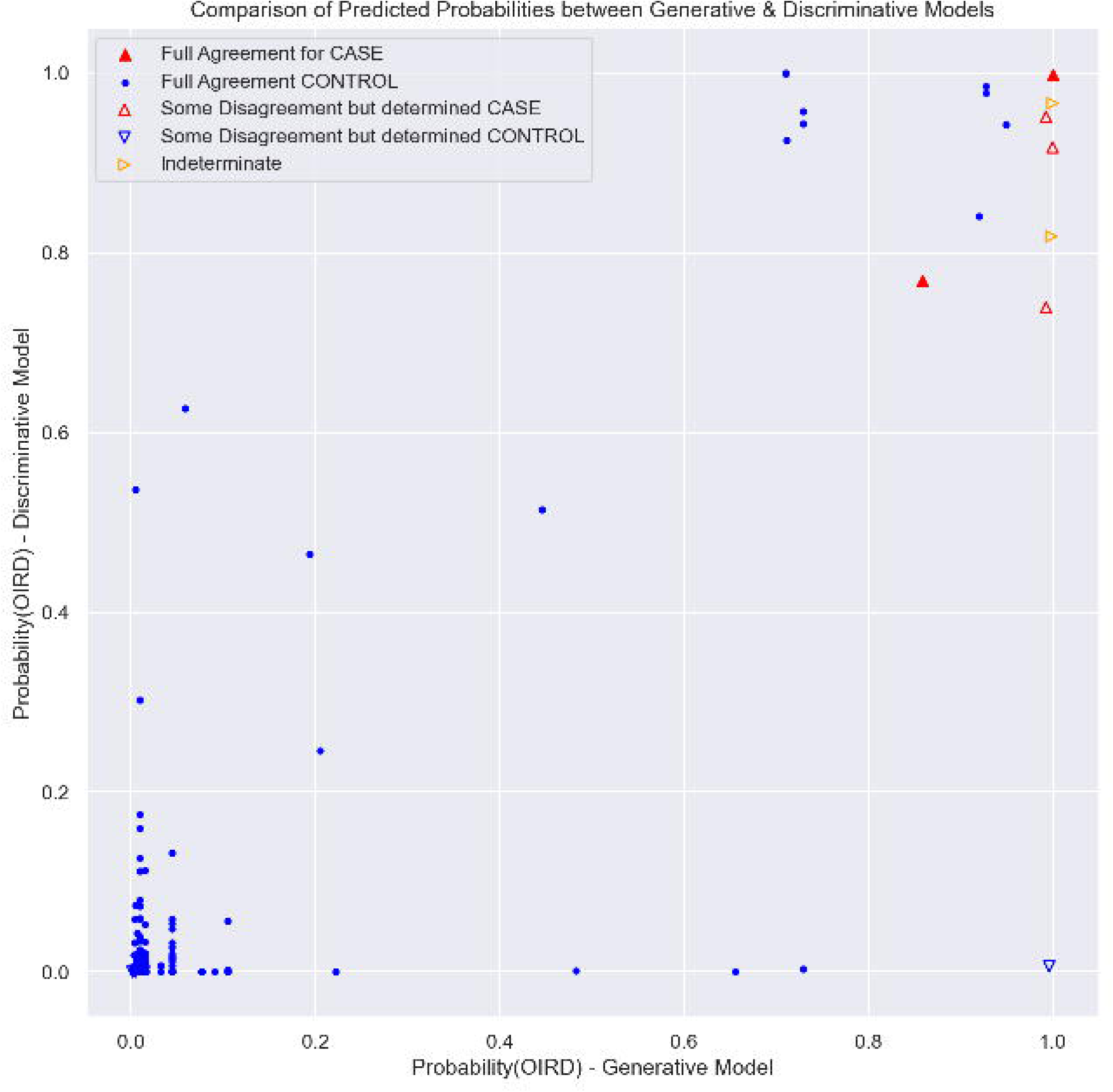
Comparison of predicted probabilities between Generative and Discriminative models with final case/control status denoted – all visits.

**Figure 6.**
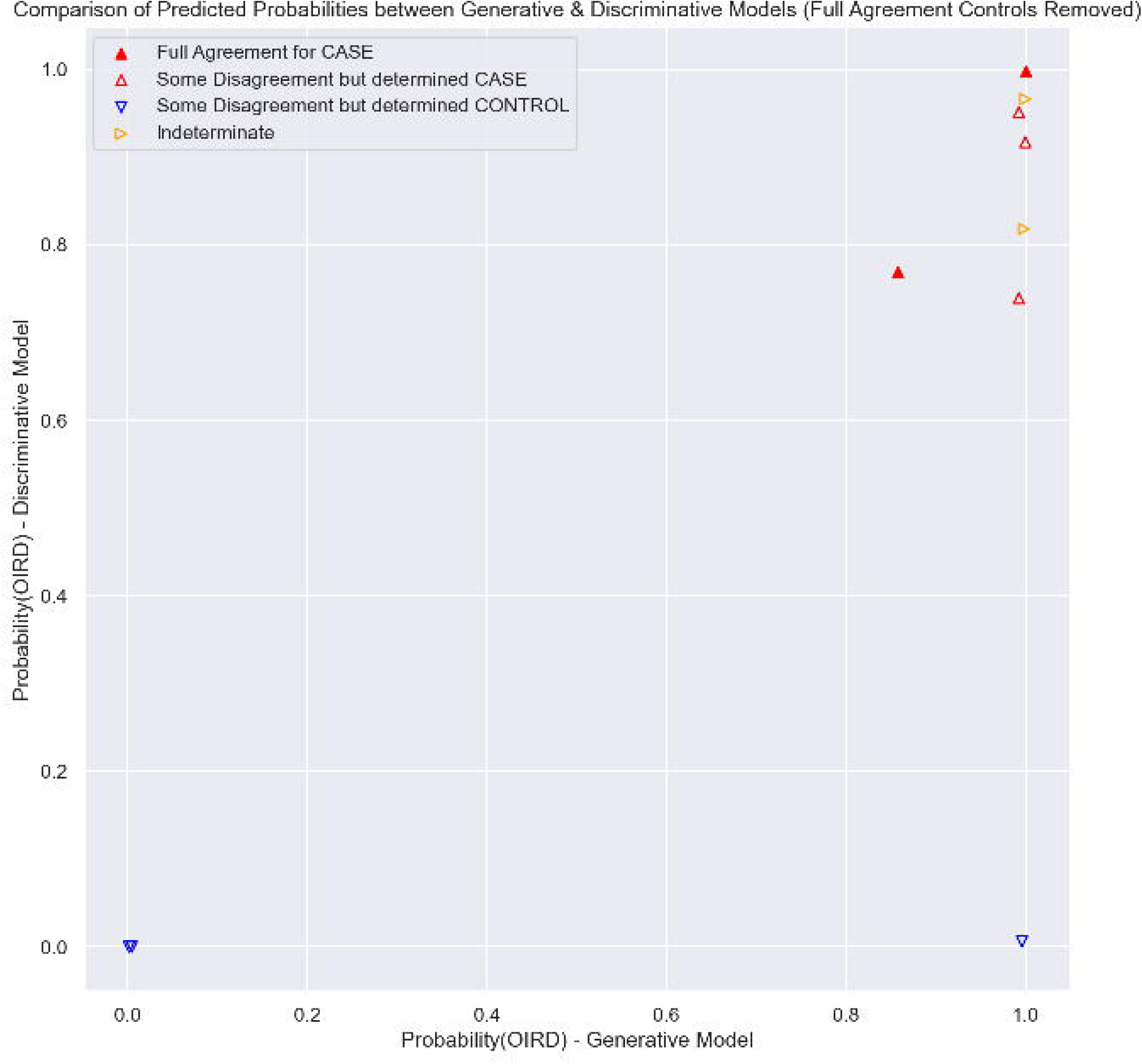
Comparison of predicted probabilities between Generative and Discriminative models with final case/control status denoted – with visits determined to be a Control with full agreement on manual review are removed.

### 3.4. review of misclassified patients

In the Validation Set, 11 of the 90 patients were classified as Cases based on the Discriminative model when the manual review classified the patients as Controls. None of the patients were misclassified as controls. Table 5 contains the predicted probabilities along with comments from manual review of the Validation Set.

**Table 5.** Predicted probabilities and manual review comments from misclassified visits in the Validation Set.

| Predicted Probability | Comments from Manual Review |
| --- | --- |
| 1.0 | Urethral cancer removal. Complex pain management for chronic cancer pain - always in pain but also became somnolent - no naloxone administration but suggested altered mental status. |
| 0.98 | Colostomy placement. Given naloxone & intubated after altered mental status - seems to be more like aspiration pneumonia. Naloxone only mentioned once & that patient became very anxious after administration. |
| 0.96 | Knee replacement. Rapid response team for respiratory compromise - likely due to metabolic acidosis or other causes. Naloxone was administered according the medication administration record but not in clinical notes (in fact one note suggests she had very little opioids). |
| 0.95 | No surgery described in clinical notes. Transferred from outside hospital for complex septicemia. Multiple notes discussed how the patient was given naloxone pre-hospital following opioid use at home. |
| 0.93 | Colectomy performed. Was somnolent & bradypneic requiring rapid response team - no effect from naloxone administration - likely due to alcohol withdrawal. |
| 0.93 | Dialysis patient presented to Emergency Department after blood cultures positive during dialysis & their operation was elective surgery for severely infected teeth. 1-month stay in the hospital. Lots of discussion regarding suboxone, high opioid use, & holding opioids. No evidence of OIRD during visit. Interestingly, the patient returned within 5 days of discharge with OIRD in the community. |
| 0.92 | Artificial hip irrigation & debridement. Coded & died after a complication with septic shock - no evidence of OIRD. Had been on oral naloxone. |
| 0.90 | No surgery described in clinical notes. Transferred from outside hospital for sepsis. Pulmonary note identified decreased respiratory drive on mechanical ventilation due to "delayed clearance of sedating meds" because they had ruled out other neurological etiologies of altered mental status. Naloxone had been administered, but this was via oral route and likely for constipation. Etiology of respiratory depression is unclear. |
| 0.82 | No surgery described in clinical notes. Transfer from outside hospital for sickle cell-related stroke. Had been taking high doses of narcotics at home. |
| 0.82 | Kidney & heart transplant. Didn't do well with extubation on post-operative day 1 & naloxone administration didn't help. Unlikely OIRD. |
| 0.78 | Pituitary tumor resection. No complications. |

During a post-hoc manual review of the Test Set visits with high (>= 0.5) Discriminative model probabilities but labeled as Controls by the crowdsourcing workers (n=14), the investigative team agreed with all crowdsourcing results and did not re-classify any Controls as Cases. However, one visit was deemed ambiguous/unclear by the crowdsourcing workers with one worker labeling the visit as a Case and one worker labeling the visit as a Control with no tie-breaker available. The investigative team re-classified the visit from Unknown to Case. Table 6 contains the predicted probabilities along with comments from the investigative team’s post-hoc manual review of the Test Set visits with high Discriminative model probabilities among Control visits.

**Table 6.**
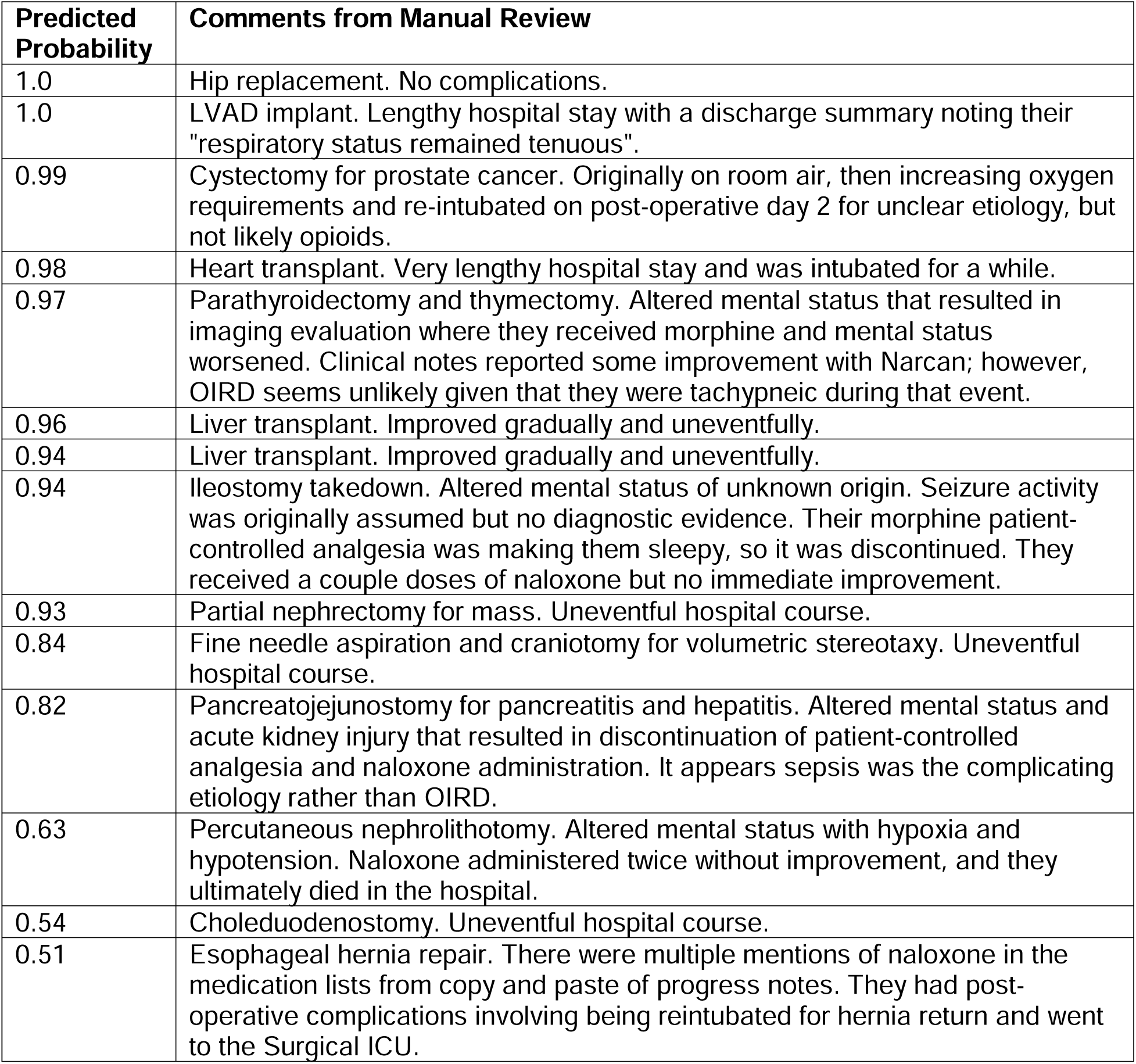
Predicted probabilities and manual review comments from misclassified visits in the Test Set.

## 4. DISCUSSION

### 4.1. summary

We applied a data programming paradigm with the use of weak learners and heterogenous data types to the problem of identifying OIRD. All manually-confirmed Cases were identified by the majority vote of LFs, Generative model, and Discriminative model. For rare outcomes like OIRD, this finding is encouraging because it can reduce the number of manual reviews needed for applying outcome labels by excluding visits/patients with low probabilities. In practice, as new patient records are added to our de-identified EHR database in the future, we could score each record with the Discriminative model quickly and follow up with a manual review only for records with high scores. While it would also be possible to use the majority vote approach or the Generative model for scoring, it is a more challenging task due to the pre-processing steps required for applying LFs and creating a label matrix.

In our post-hoc sensitivity analysis of potential information added to the Discriminative model in the Validation Set, our results suggested sample weighting (based on the degree of uncertainty in the Generative model) improved overall performance and incorporating the outcome labels from manual adjudication corrected some misclassification. This latter finding is likely due to the iterative enrichment of our Development Set and Validation Set with the top 20 Generative model probabilities as we developed LFs. Enriching both Sets with relatively homogenous records (i.e., the highest probabilities) and then building a Discriminative model with the combined Training and Development Sets resulted in added information that improved predictions in the Validation set. We did not find this added information influenced performance in the hold-out Test Set where the Generative and Discriminative models performed similarly. However, we did observe improved performance in the Test Set of the unweighted model as well as removal of the manually adjudicated labels. This observation suggests our final Discriminative model was slightly over-fit with a higher number of false positives. To advance the science of computational phenotyping, future studies should continue examining which modeling choices are ideal for certain scenarios and assumptions.

Other biomedical studies have used the paradigm proposed by Snorkel (e.g., post-market medical device surveillance[27], extraction of pain levels from EHR notes[15]). Others’ work using Snorkel suggests the Discriminative models perform better than Generative models,[15] so we hypothesized model performance on the hold-out Test Set would be high. What we found was that the two models performed differently, and there could be merit in considering both for creating outcome labels.

### 4.2. limitations

Our work has its limitations. That study was conducted in a single organization, which could limit generalizability. Our data source did not identify the elective nature of its surgeries, which we attempted to overcome with the removal of visits where the surgical date occurred on the same day as an Emergency Department visit. Another limitation of our work is a relative reduction in the potential data types included in LFs. For example, when exploring the effectiveness of naloxone administration, we attempted to incorporate the cosine similarity of vector embeddings of text data compared to examples of text suggesting naloxone effectiveness without success. Future studies could examine whether this contemporary natural language processing method improves LF performance. Similarly, clinical notes authored by nurses were not typically available in our data source. Although it is unlikely a nurse would document evidence of OIRD when a prescribing provider does not, that scenario could occur and should be examined in future work.

We initially followed the Snorkel developers’ guidance for all steps in the labeling process but ultimately made some modifications, which we believe add to the literature for computational phenotyping of health-related conditions. During hyperparameter tuning of the Generative model, we used a single reviewer to determine which LF rank ordering had the greatest face validity for clinical relevance. Additional work is needed to explore whether a more reliable and valid approach for determining the most appropriate ranking is possible, particularly as this was a departure from using Snorkel’s recommendation of empirical accuracy. Finally, our iterative LF development process depended on enriching the Development Set and Validation Set based on the highest probabilities of candidate Generative models. We did not enrich our data sets for Control status (i.e., lower probabilities), but Control enrichment could easily be included depending on the clinical outcome under investigation.

## 5. CONCLUSION

The use of Snorkel to implement a data programming approach for phenotyping OIRD in a large observational data set was successful, particularly with its 100% sensitivity. This method opens new opportunities for identifying rare, incompletely ascertainable outcomes in large clinical data sets. Although the F1 score suggested only moderate overall performance, the high sensitivity of Snorkel’s predictions combined with the low prevalence of OIRD results in significantly fewer manual chart reviews (compared to not using Snorkel) necessary to apply phenotypes to the entirety of a large data set. In the future, we plan to apply Snorkel to other clinical domains to evaluate performance and explore under what conditions (e.g., data types, data quality, number of labeling functions, scientific programming experience of research investigators) Snorkel performs well.

## Declarations

## Funding Acknowledgements

We received support for this work from the Agency for Healthcare Research & Quality (AHRQ) and the Patient-Centered Outcomes Research Institute (PCORI) under Award Number K12 HS026395; resources and use of facilities at the Department of Veterans Affairs, Tennessee Valley Healthcare System, in collaboration with the Medical Informatics Fellowship; the Vanderbilt Institute for Clinical and Translational Research (VICTR) under Award Number UL1 TR000445 from NIH/NCATS; and the Advanced Computing Center for Research and Education (ACCRE) High-Memory Compute Nodes under Grant# 1S10OD023680-01. The content is solely the responsibility of the authors and does not necessarily represent the official views of AHRQ, PCORI, the NIH, the Department of Veterans Affairs, or the United States Government.

## Data Availability Statement

The data underlying this article cannot be shared publicly in order to protect the privacy of the individuals whose medical records we used. Data can be made available with a written request to the corresponding author.

## Ethics Statement

We received Institutional Review Board approval from Vanderbilt University Medical Center under approval numbers 201918 and 171618.

## Notes

### Competing Interest Statement

The authors have declared no competing interest.

### Author Declarations

The IRB of Vanderbilt University Medical Center ethical approval for this work under studies #171618 and #201918.

